# Impact of Social Distancing Measures on COVID-19 Healthcare Demand in Central Texas

**DOI:** 10.1101/2020.04.16.20068403

**Authors:** Xutong Wang, Remy F Pasco, Zhanwei Du, Michaela Petty, Spencer J Fox, Alison P Galvani, Michael Pignone, S. Claiborne Johnston, Lauren Ancel Meyers

**Author notes:** Contributed equally (Xutong Wang, Remy F Pasco, Zhanwei Du).

## Abstract

**Background:** A novel coronavirus (SARS-CoV-2) emerged in Wuhan, China in late 2019 and rapidly spread worldwide. In the absence of effective antiviral drugs and vaccines, well-targeted social distancing measures are essential for mitigating the COVID-19 pandemic, reducing strain on local health systems, and preventing mortality. Here, we provide a quantitative assessment of the efficacy of social distancing to slow COVID-19 transmission and reduce hospital surge, depending on the timing and extent of the measures imposed for a metropolitan region and its health care systems.

**Methods and Findings:** We built a granular mathematical model of COVID-19 transmission that incorporated age-specific and risk-stratified heterogeneity, estimates for the transmission, and severity of COVID-19 using current best evidence. We performed thousands of stochastic simulations of COVID-19 transmission in the Austin-Round Rock Metropolitan Area to project the impact of school closures coupled with social distancing measures that were estimated to reduce non-household contacts by 0%, 25%, 50%, 75% or 90%. We compare early versus late implementation and estimate the number of COVID-19 hospitalizations, ICU patients, ventilator needs and deaths through mid-August, 2020. We queried local emergency services and hospital systems to estimate total hospital bed, ICU, and ventilator capacity for the region. We expected COVID-19 hospital beds and ICU requirements would surpass local capacity by mid-May if no intervention was taken.

Assuming a four-day epidemic doubling time, school closures alone would be expected to reduce peak hospitalizations by only 18% and cumulative deaths by less than 3%. Immediate social distancing measures that reduced non-household contacts by over 75%, such as stay-at-home orders and closing of non-essential businesses, would be required to ensure that COVID-19 cases do not overwhelm local hospital surge capacity. Peak ICU bed demand prior to mid August 2020 would be expected to be reduced from 2,121 (95% CI: 2,018-2,208) with no intervention to 698 (95% CI: 204-1,100) with 75% social distancing and 136 (95% CI: 38-308) with 90% social distancing; current ICU bed capacity was estimated at 680. A two-week delay in implementation of such measures is projected to accelerate a local ICU bed shortage by four weeks.

**Conclusions:** School closures alone hardly impact the epidemic curve. Immediate social distancing measures that reduce non-household contacts by over 75% were required to ensure that COVID-19 cases do not overwhelm local hospital surge capacity. These findings helped inform the Stay Home-Work Safe order enacted by the city of Austin, Texas on March 24, 2020 as a means of mitigating the emerging COVID-19 epidemic.

## Introduction

A novel coronavirus (SARS-CoV-2) appeared in Wuhan, China in December 2019 and was declared a pandemic on March 11, 2020 by the World Health Organization (WHO) [1]. As of March 28, 2020, 191 countries, areas, or territories have reported a total of 571,659 confirmed COVID-19 cases and 26,493 deaths, with substantial outbreaks emerging in Italy, the Republic of Korea, Iran, and the United States, which surpassed China to become the country with the highest cumulative confirmed cases [2,3].

The US reported its first imported SARS-CoV-2 case from Wuhan on January 20, 2020 in Washington state [4], 6 and 40 days ahead of California and New York City [5,6], and its first locally-infected cases on February 28, 2020 [7]. The number of confirmed cases has continued to grow exponentially [8]. As of March 26, 2020, all 50 states have reported confirmed cases, 27 have reported community spread and the cumulative confirmed COVID-19 cases are 68,440 with 994 total deaths [8]. Surges in COVID-19 hospitalizations are undermining local healthcare systems in New York and Seattle [9,10].

In order to contain the spread of SARS-CoV-2, cities have implemented social distancing measures at multiple scales, including school closures, limiting mass gatherings, issuing stay at home orders, imposing travel restrictions, and banning non-essential commercial activities. As of March 28, 2020, 26 states have issued a statewide shelter-in-place order or have at least one city-level stay-at-home order, affecting over 229 million people [11]. However, the timing of interventions has been idiosyncratic and many cities have yet to enact substantial social distancing orders [12,13].

As COVID-19 emerged into a global threat, we took a national pandemic influenza model that was built through a pandemic preparedness contract with the CDC and adapted it to model the spread and control of COVID-19 within and between 217 US cities. We used this model to project the potential impact of school closures coupled with social distancing, in terms of reducing cases, deaths, hospitalizations, ICU visits, and ventilator needs, on local, regional and national scales. Here, we have focused our analysis on the city of Austin, which is the capital of Texas and the fastest growing city in the US [14,15]. as a representation of major US metropolitan areas. The scenarios and inputs (e.g., epidemiological parameters) were determined in consultation with the US Centers for Disease Control and Prevention and the city of Regional Healthcare System Executive Council of the Austin-Travis County Emergency Operations Command.

## Methods

We focused on the Austin-Round Rock Metropolitan Area with a population of 2.17 million people in 2018, but conjecture that the qualitative findings and impact of social distancing will apply to cities throughout the US. We analyzed a compartmental model that incorporates age-specific high risk proportions and contact rates to measure the impact of two key interventions: (1) school closures and (2) social distancing measures that reduce non-household contacts by a specified percent. We estimated the impact of these measures on cases, hospitalizations, ICU visits, ventilator needs, and deaths.

### Transmission model

We built a stochastic age- and risk-structured susceptible-exposed-asymptomatic-symptomatic-hospitalized-recovered (SEAYHR) model of SARS-CoV-2 transmission (Fig. S1). Individuals were separated into five age groups: 0-4, 5-7, 18-49, 50-64, and 65+ years old based on population data for the five-county Austin-Round Rock Metropolitan Area from the 2017 American Community Survey [16]. Each age group was divided into a low-risk and high-risk group, based on the prevalence of chronic conditions estimated for the Austin population (Fig. S2) [17–20]. We also estimated the proportion of pregnant women in each age group as a special risk class [21]. All individuals were assumed to be susceptible to the disease. Infected individuals were modeled to enter an incubation period where they were symptom-free but could be mildly infectious [22] and then progressed to either a symptomatic or asymptomatic compartment. Asymptomatic individuals were assumed to have the same infectious period as symptomatic individuals but lower infectiousness. The rates at which symptomatic cases were moved to a hospitalized compartment and died depended on both age and risk group. Recovered individuals were considered fully immune. The Supplement describes the methods in greater detail.

All model parameters are provided in Tables S1.1-S1.3 and were based on published estimates from COVID-19 studies as well as input from the US CDC and City of Austin. We assumed a basic reproduction number (*R*_0_) of 2.2 [23] and considered two different doubling times of 7.2 days (low growth rate) [23–26] and 4 days (high growth rate) [25,27,28]. Age-specific contact rates were estimated using contact matrices published by Prem et al. and are adjusted to model school closures and various levels of social distancing [29]. Transmission rates were estimated by fitting simulations to a given *R*_0_ and epidemic doubling time. The incubation period was sampled from a triangular distribution from 5.6 days to 8.2 days with mean of 7 days [30] and the infectious period was sampled from a triangular distribution from 21.1 days to 24.4 days with mean of 22.6 days [31]. We assumed the asymptomatic ratio to be 17.9% [32] with 12.6% of infections arising from pre-symptomatic transmission during the incubation period [22]. Following the CDC’s planning scenarios, we assumed that the infection hospitalization rate and infection fatality rate was ten times higher in high-risk than low risk individuals, within each age group.

Simulations began with five imported symptomatic cases in the 18-49 year-old age group on March 1, 2020 and update at 2.4-hour intervals. For each combination of epidemic scenarios (low / high growth rate) and intervention strategies (school closure policy with different levels of social distancing), we ran 100 stochastic simulations and reported the medians and 95% range at weekly intervals.

### School closure policies

As part of a CDC modeling network, we initially modeled a large number of school closure policies, with variable implementation time and duration. Here, we reported only two of these strategies to demonstrate the impact of implementation time: (i) closure immediately following the first confirmed case (March 14) and (ii) delayed closure two months after the first confirmed case (May 14). In both cases we assumed that schools remain closed through the end of the summer vacation (August 18, 2020) [33], which corresponds to a 23-week duration for the early closure, and a 14-week duration for the late closure. The early closure scenario roughly corresponded to the city of Austin announcing the first two confirmed cases on March 13th and major school districts closing the next day. In our simulations, the median cumulative number of symptomatic COVID-19 cases by March 14th was 23 (IQR: 17-32) and 9.0 (IQR: 5.8-13) assuming a four-day and seven-day doubling period, respectively; by May 14th, median cumulative symptomatic cases climbed to 508,546 (IQR: 104,535-766,109) and 6,759 (IQR: 1,901-12,353), respectively.

### Social distancing measures

In addition to school closures, we considered the effect of various levels of social distancing that decrease non-household contacts by 25%, 50%, 75%, and 90% overall. These levels were chosen to correspond to increasingly more severe levels of restriction on social interaction from limiting large crowds to near-total restriction on out of home movement except for health care and basic necessities.

Age-stratified contact rates [29] were derived from the POLYMOD diary-based study in Europe [34] and separated in contacts occurring at home, at school, at work and elsewhere. We used the national US age distribution[35] to aggregate these estimates from 17 to the five age groups of our model (Tables S1.4-S1.7). We combined these matrices to model four different types of days: (i) normal school days (all contacts), (ii) normal weekends and short weekday holidays (all but school and work contacts; adults are assumed to work during the long summer break), (iii) weekdays during school closures/social distancing, (iv) weekend or weekday holiday during school closure/social distancing. To model school closures with social distancing, we included all household contacts plus a specified proportion of contacts outside the home. On weekdays, this included a proportion of contacts occurring at work and elsewhere; on weekends and holidays (excluding summer vacation) this included just contacts occurring elsewhere. Days were assigned to one of these four contact models based on the 2019-2020 and 2020-2021 school calendars from the Austin Independent School District [36], which was the largest public school district in the metropolitan area serving approximately 22.7% of the Austin-Round Rock MSA population.

### Healthcare demands

We assumed that hospitalized cases were admitted on average 5.9 days [37] following symptom onset, with the infection hospitalization rate depending on the age and risk group (Table S1.1). Hospitalized cases who recovered were considered discharged an average of 11.5 days following admission [38]; deaths occured an average of 11.2 days following admission [38]. We estimated the number ICU beds and ventilators needed to care for COVID-19 cases each day based on age-specific rates provided by the CDC (Table S1.3) and assuming that the average duration of ICU care and ventilation support are 8 days and 5 days, respectively. There is some uncertainty regarding how these estimates may change when healthcare facilities reach or exceed capacity, because of a lack of available post-discharge care and inefficiency in the healthcare system due to worker illness. Thus, we also tested an alternative scenario with significantly longer duration of both ICU care and ventilation (SI S3). We did not consider potential excess mortality resulting from lack of access to adequate healthcare during pandemic surges.

## Results

Our analyses focus on two key *levers* of intervention––the speed of implementation and the extent of social distancing. We consider two scenarios for the epidemic growth rate of COVID-19 and project five *outcomes*––cases, hospitalizations, ICU care, ventilator needs and deaths.

Regardless of epidemic growth rate, school closures alone had little effect on the speed and burden of the epidemic (Fig. 1). High levels of social distancing, when coupled with school closures, substantially delayed and dampened the epidemic peak. The impact of the measures depended on early implementation. Under both the slower and faster epidemic growth scenarios (i.e., seven-day and four-day doubling times), immediate measures beginning on March 14th were much more effective than two-month delayed measures at slowing transmission throughout the spring and summer of 2020 (Fig. 1). Given that recent estimates for the doubling time in US cities are quite short, ranging from 2.4 to 3 days [28,39], this suggests that delayed measures will be almost entirely ineffective.

**Fig 1.**
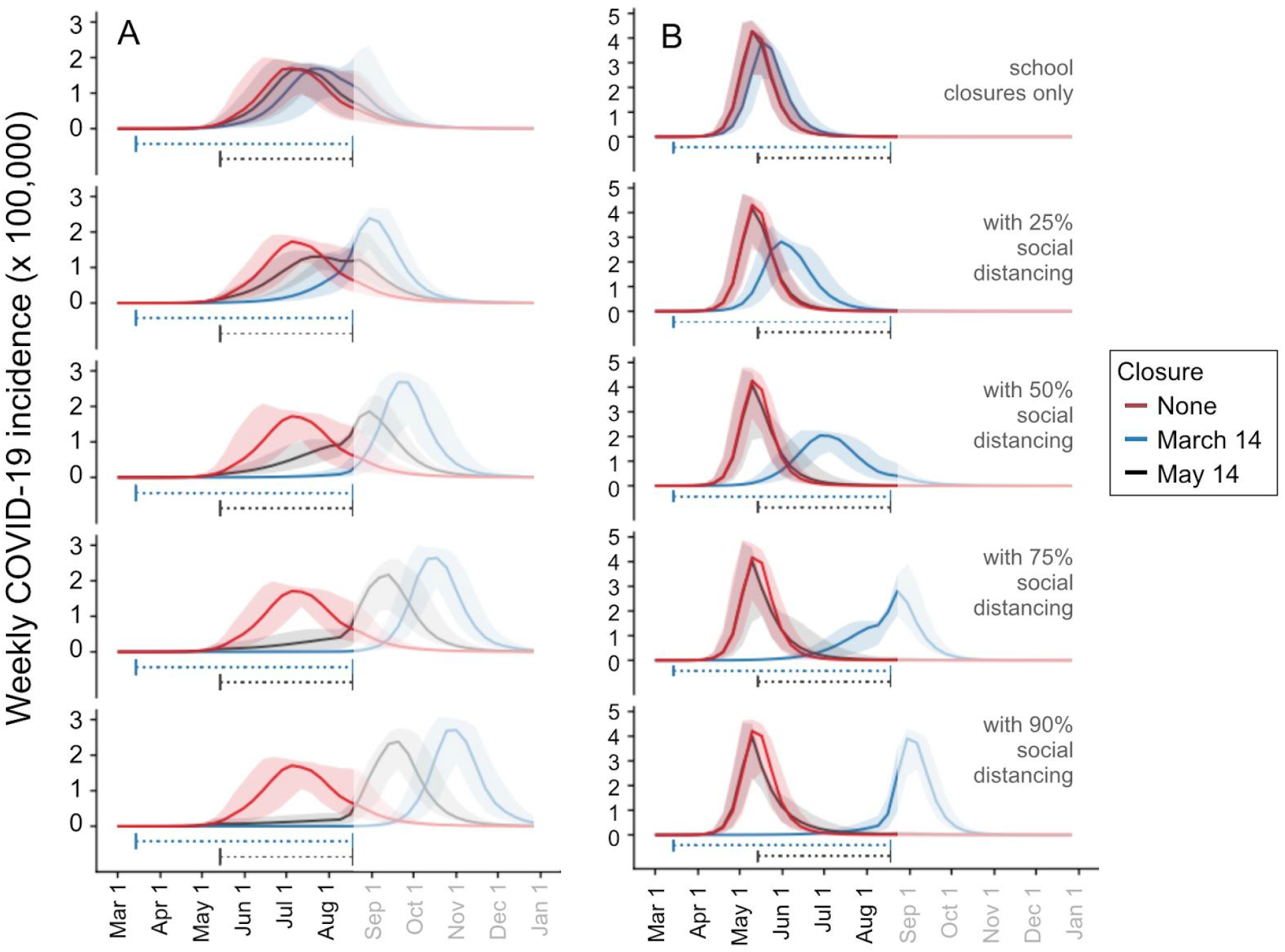
Projected weekly incident COVID-19 cases in the Austin-Round Rock MSA. Graphs show simulation results for different levels of social distancing and implementation times, assuming an epidemic doubling time of (A) 7.2 days [22–25] or (B) 4 days [25,27,28]. Each graph displays three projections: a baseline assuming no social distancing (red), social distancing implemented March 14-Aug 17, 2020 (blue), and social distancing implemented May 14-Aug 17, 2020 (black). From top to bottom, the graphs in each column correspond to increasingly stringent social distancing measures: school closures plus social distancing that reduces non-household contacts by 0%, 25%, 50%, 75%, or 90%. Solid lines indicate the median of 100 stochastic simulations; shading indicates the inner 95% range of values. The horizontal dotted lines beneath the curves indicate intervention periods. The faded mid-August to December time range indicates long-range uncertainty regarding COVID-19 transmission dynamics and intervention policies.

To assess the impact of social distancing measures on mitigating healthcare surge in the Austin-Round Rock MSA, we considered the more plausible four-day doubling time scenario (Table 1 and Fig. 2). Social distancing measures that reduced non-household contacts by less than 75% were projected to delay but not prevent a healthcare crisis. Only the 90% contact reduction scenario was projected to reduce hospitalizations, ICU care and ventilator needs below the estimated capacity for the metropolitan area (Table 2). If 75% social distancing were implemented on March 28th instead of March 14th (i.e., a two-week delay), we would expect COVID-19 ICU requirements to exceed local capacity by mid-July instead of mid-August (i.e., a four-week acceleration) (SI S4).

**Table 1.** Estimated cumulative COVID-19 cases, healthcare requirements and deaths. The values are medians across 100 stochastic simulations for the Austin-Round Rock MSA from March 1 through August 17, 2020 based on the parameters given in Table S1.

| **Outcomes** | **No measures** | **School closure** | **School closure + 50% social distancing** | **School closure + 75% social distancing** | **School closure + 90% social distancing** |
| --- | --- | --- | --- | --- | --- |
| **Cases** | 1,766,480 | 1,761,426 | 1,563,654 | 698,640 | 139,589 |
| **Hospitalizations** | 87,770 | 86,720 | 61,620 | 18,309 | 3,432 |
| **ICU** | 14,704 | 14,528 | 10,344 | 3,082 | 578 |
| **Ventilators** | 6,919 | 6,836 | 4,870 | 1,452 | 272 |
| **Deaths** | 10,961 | 10,660 | 6,245 | 1,508 | 276 |

**Table 2.**
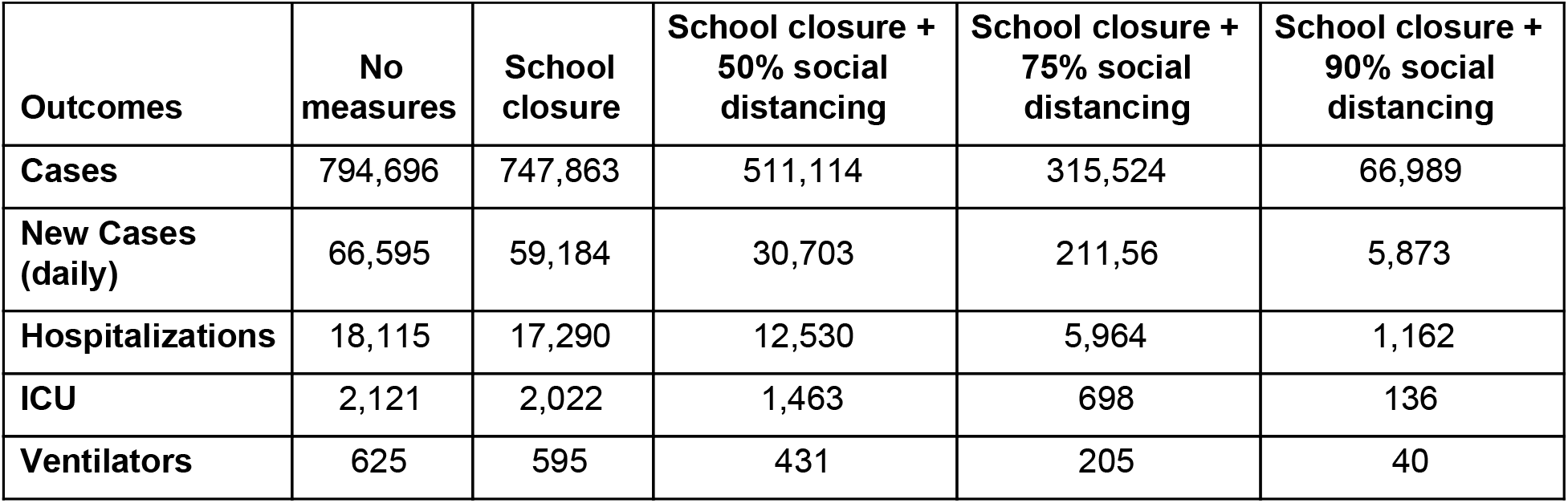
Estimated peak COVID-19 cases and healthcare demands. The values are medians across 100 stochastic simulations for the Austin-Round Rock MSA from March 1 through August 17, 2020 based on the parameters given in Table S1.

| **Outcomes** | **No measures** | **School closure** | **School closure + 50% social distancing** | **School closure + 75% social distancing** | **School closure + 90% social distancing** |
| --- | --- | --- | --- | --- | --- |
| **Cases** | 794,696 | 747,863 | 511,114 | 315,524 | 66,989 |
| **New Cases (daily)** | 66,595 | 59,184 | 30,703 | 211,56 | 5,873 |
| **Hospitalizations** | 18,115 | 17,290 | 12,530 | 5,964 | 1,162 |
| **ICU** | 2,121 | 2,022 | 1,463 | 698 | 136 |
| **Ventilators** | 625 | 595 | 431 | 205 | 40 |

**Fig 2.**
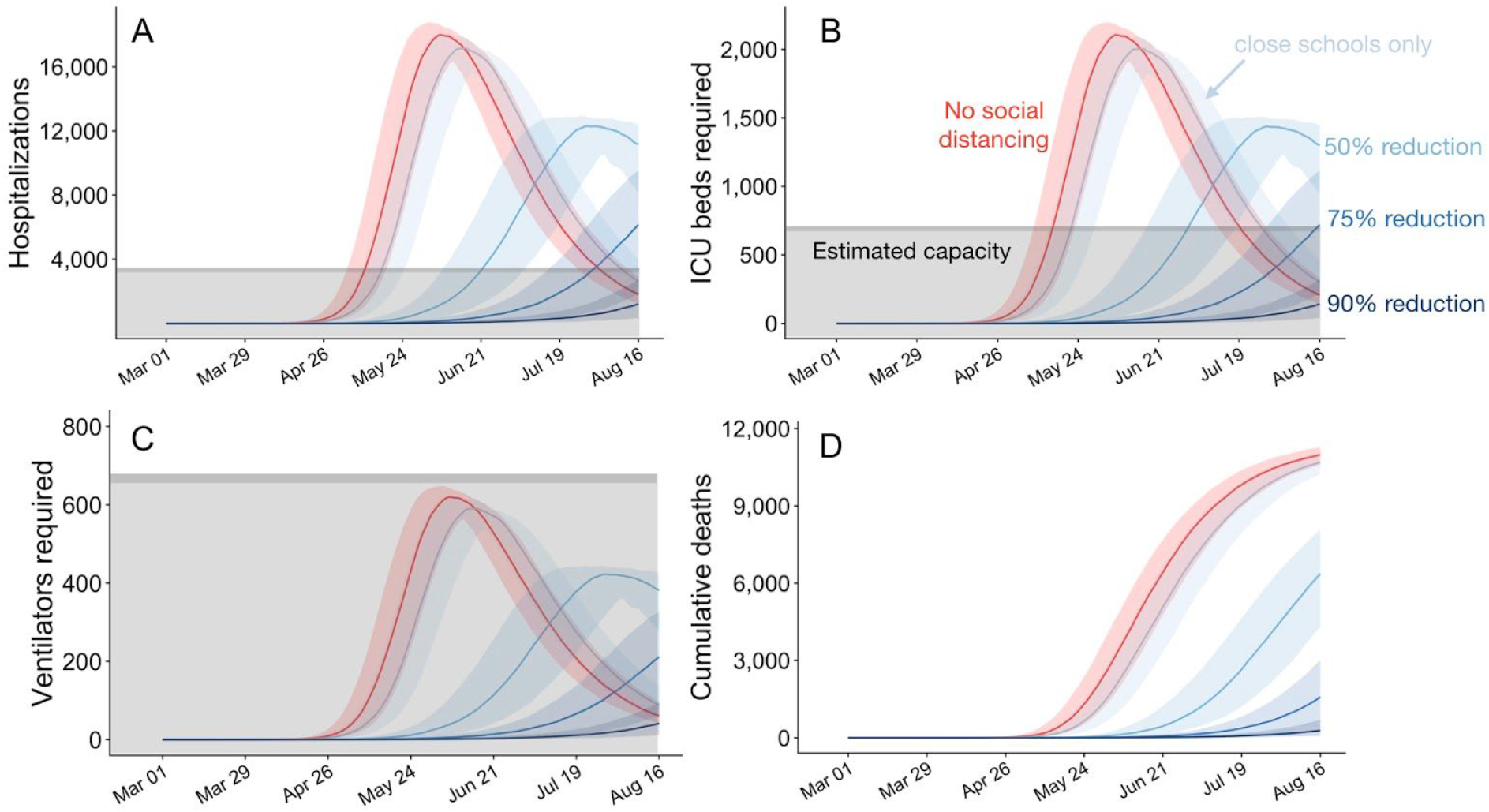
Projected COVID-19 healthcare demand and cumulative deaths in the Austin-Round Rock MSA from March 1 to August 17, 2020. Graphs show simulation results across multiple levels of social distancing, assuming *R*_0_=2.2 with a four-day epidemic doubling time. Extensive social distancing is expected to substantially reduce the burden of COVID-19 (A) hospitalizations, (B) patients requiring ICU care, (C) patients requiring mechanical ventilation and (D) mortality. The red lines project COVID-19 transmission assuming no interventions under the parameters given in Table S1. The blue lines show increasing levels of social distancing interventions, from light to dark: school closures plus social distancing interventions that reduce non-household contacts by either 0%, 50%, 75% or 90%. Lines and shading indicate the median and inner 95% range of values across 100 stochastic simulations. Gray shaded region indicates estimated surge capacity for COVID-19 patients in the Austin-Round Rock MSA as of March 28, 2020, which is calculated based on 80% of the total 4299 hospital beds and 90% of the total 755 ICU beds and 755 mechanical ventilators.

Under the naive scenario that school closures and social distancing measures are lifted *entirely* on the first day of the 2020-2021 academic year (August 18) [33], *the pace and* extent of COVID-19 transmission in the fall would depend on how many people were infected (and thereby immunized) during the spring and summer (Fig. 1). As cumulative incidence approaches the herd immunity threshold of roughly 55% of the population, the reproduction number *(R*_0_) declines. Once this 55% threshold is surpassed, the reproduction number falls below one and the virus would be unable to spread widely, even if social distancing measures are lifted. Assuming the faster four-day epidemic doubling time (Fig. 1B), only the highest levels of social distancing suppress transmission over the summer. Under 75% or 90% social distancing, the lifting of measures on August 18th would be expected to produce epidemic peaks in the first or second week of September, respectively. Assuming the slower seven-day doubling time (Fig. 1A), even delayed social distancing would be expected to forestall the start of the epidemic from spring to fall. The higher fall peaks that were produced under the most extreme social distancing stem from baseline contact patterns (in the absence of social distancing): a COVID-19 epidemic that begins in the spring would be naturally dampened by the three month summer vacation period when children are out of school, whereas fall start would be amplified by the start of the academic year.

## Discussion

As COVID-19 emerged as a global threat in early 2020, we rapidly adapted a pandemic influenza model that was under development as part of an effort coordinated by the US CDC to build a strategic national modeling resource for pandemic planning and response. The analyses presented herein originated in time-sensitive requests from the CDC, the city of Austin, and the state of Texas to evaluate the potential impact of school closures and social distancing on the emergence and spread of COVID-19 in US cities. Our projections indicate that without extensive social distancing measures, the emerging outbreak will quickly surpass healthcare capacity in the region. However, with extensive social distancing, the number of cases, hospitalizations, and deaths can be substantially reduced throughout the summer of 2020. Although these analyses are specific to the Austin-Round Rock metropolitan area, we expect that the impacts of the mitigation strategies will be qualitatively similar for cities throughout the US.

Our epidemiological projections and conclusions regarding the urgent need for extensive social distancing are consistent with a recent analysis by Imperial College [40]. However, we assume that a lower percent of hospitalized patients receive critical care (15-20% versus 30%), and consequently project a lower peak ICU demand. In sensitivity analyses with more extreme assumptions about critical care requirements, the projected peak demand rises accordingly. The local focus of our model, which incorporates city-specific data regarding demographics, high-risk conditions, contact patterns, and healthcare resource availability, allows us to project near-term healthcare demands and provide actionable insights for local healthcare and governmental decision-makers.

We conducted these analyses to inform decision making in a rapidly evolving environment with substantial uncertainty. On March 6, 2020, the city of Austin declared a local state of disaster and cancelled the South by Southwest Conference and Festival (SXSW) which was expected to draw 417,400 visitors from around the world and bring $355.9 million to the local economy [41]. Evidence of community transmission appeared within days of the first confirmed COVID-19 case in Austin, on March 13, 2020. Shortly after, the University of Texas at Austin, one of the largest public universities with over 50,000 students [42]. and the largest public school district in Austin announced school closures [43,44]. On March 24, 2020, the city of Austin issued a ‘Stay Home – Work Safe’ order to eliminate all non-essential business and travels [45]. Austin’s City leaders requested the healthcare analyses presented in Figure 2 in the days leading up to the March 24th order and requested that we release a preliminary report to educate the public [46,47].

Social distancing measures, including school closures, restrictions on travel, mass gatherings and commercial activities, and more extensive shelter-in-place advisories, aim to decrease disease transmission within a population by preventing contacts between people. Our analyses project the impacts of such measures on the transmission dynamics of COVID-19, but do not consider the economic, social and psychological costs of social distancing measures, including the socioeconomic disparities in burden and morbidity and mortality resulting from reductions in health and mental health care services [48,49].

There is an urgent need to project the relative impact of different levers for social distancing in light of their potential societal costs––including school closures, partial work and travel restrictions and cocooning of the high risk––so that restrictions can be strategically lifted without compromising public health. In particular, school closures are often deployed earlier than more extensive social distancing measures. Yet, they can be costly, particularly for low-income families who may rely on lunch programs and be unable to afford childcare [50,51], and our analysis suggests that they may only slightly reduce the pace of transmission and peak hospital surge. The role of children in community transmission of COVID-19 remains uncertain; thus, school closures are prudent at this time. Children represent a low proportion or confirmed cases worldwide [31,52], perhaps reflecting that COVID-19 is less severe in children than adults [53,54]. If we learn that the prevalence or infectiousness of COVID-19 is low in children, then opening schools may be a reasonable first step towards resuming normalcy.

Although our model incorporates considerable detail regarding the natural history of COVID-19, age- and location-specific contact patterns, and the demographic and risk composition of the Austin-Round Rock MSA, it does not explicitly capture neighborhood, household or other community structure that can serve to amplify or impede transmission [55–57]. In addition, we ignore the possible importation of COVID-19 cases from other cities, under the assumption that the additional cases will have a negligible impact particularly during the period of exponential growth. Our model also does not evaluate other potentially effective interventions such as increased levels of selective testing and isolation.

These analyses rely on recently published estimates for transmission rate and severity of COVID-19 as well as best estimates from expert opinions from the CDC and Dell Medical School. There is still much we do not understand about the transmission dynamics of this virus, including the extent of asymptomatic infection and transmission. Given that our understanding of COVID-19 is evolving so rapidly, we expect that there may be consensus around different estimates for key transmission and severity parameters by the time this work is published. Thus, we emphasize the qualitative but not quantitative results of the analysis.

Given the rapid spread of COVID-19, early and extensive social distancing are both viable and necessary for preventing catastrophic hospital surges. Despite uncertainties in key parameters and the focus on a single city, the expansion and containment of COVID-19 in cities worldwide suggest that these insights are widely applicable. This framework can be updated as situational awareness of COVID-19 improves to provide a quantitative sounding board as public health agencies evaluate strategies for mitigating risks while sustaining economic activity in the US.

## Data Availability

Not applicable

## Acknowledgments

We acknowledge the critical discussions and parameter guidance from Matthew Biggerstaff, Michael Johannson, and the FluCode network at the US CDC, Mayor Steven Adler of Austin, Texas, and the White House Coronavirus Task Force.

